# Antiviral activities of sotrovimab against BQ.1.1 and XBB.1.5 in sera of treated patients

**DOI:** 10.1101/2023.05.25.23290512

**Authors:** Timothée Bruel, Lou-Léna Vrignaud, Françoise Porrot, Isabelle Staropoli, Delphine Planas, Florence Guivel-Benhassine, Julien Puech, Matthieu Prot, Sandie Munier, William Henry-Bolland, Cathia Soulié, Karen Zafilaza, Clovis Lusivika-Nzinga, Marie-Laure Meledge, Céline Dorival, Diana Molino, Hélène Péré, Youri Yordanov, Etienne Simon-Lorière, David Veyer, Fabrice Carrat, Olivier Schwartz, Anne-Geneviève Marcelin, Guillaume Martin-Blondel, the ANRS 0003S CoCoPrev Study Group

**Author notes:** Correspondence to &. Equal contributions (Olivier Schwartz, Anne-Geneviève Marcelin, Guillaume Martin-Blondel).

## Abstract

**Background:** Monoclonal antibodies (mAbs) targeting the spike of SARS-CoV-2 prevent severe COVID-19. Omicron subvariants BQ.1.1 and XBB.1.5 evade neutralization of therapeutic mAbs, leading to recommendations against their use. Yet, the antiviral activities of mAbs in treated patients remain ill-defined.

**Methods:** We investigated neutralization and antibody-dependent cellular cytotoxicity (ADCC) of D614G, BQ.1.1 and XBB.1.5 in 320 sera from 80 immunocompromised patients with mild-to-moderate COVID-19 prospectively treated with mAbs (sotrovimab, n=29; imdevimab/casirivimab, n=34; cilgavimab/tixagevimab, n=4) or anti-protease (nirmatrelvir/ritonavir, n=13). We measured live-virus neutralization titers and quantified ADCC with a reporter assay.

**Findings:** Only Sotrovimab elicits serum neutralization and ADCC against BQ.1.1 and XBB.1.5. As compared to D614G, sotrovimab neutralization titers of BQ.1.1 and XBB.1.5 are reduced (71- and 58-fold, respectively), but ADCC levels are only slightly decreased (1.4- and 1-fold, for BQ.1.1 and XBB.1.5, respectively).

**Interpretation:** Our results show that sotrovimab is active against BQ.1.1 and XBB.1.5 in treated individuals, suggesting that it may be a valuable therapeutic option.

## Introduction

Antiviral agents including anti-spike monoclonal antibodies (mAbs) and small molecules targeting viral enzymes remain of outmost importance to protect the most vulnerable against severe COVID-19^1^. Therapeutic mAbs (Bamlanivimab, Etesivimab, Casirivimab, Imdevimab, Cilgavimab, Tixagevimab, Bebtelovimab and Sotrovimab) target the receptor binding domain (RBD) on the viral spike (S) protein. They inhibit viral entry in a process called neutralization^2^. mAbs reduce the risk of COVID-19-related hospitalization and death in patients who are at high-risk of progression to severe COVID-19^3–5^. However, the emergence of viral variants led to a reduction of their efficacy. This climaxed with the omicron subvariants BQ.1.1 and XBB.1.5, which fully or partially resist neutralization by all available therapeutic mAbs^6,7^. Therefore, as of May 2023, mAbs are no longer recommended as a first-line treatment of COVID-19^8^. Small molecules such as nirmaltrevir, an inhibitor of the viral 3C-like protease, or remdesivir and molnupinavir, two nucleoside analogs with broad-spectrum antiviral activities, also reduce, to various extents, the progression to severe COVID-19^9–11^. Their efficacy is conserved across SARS-CoV-2 variants^7^, but their use their route of administration or their drug-drug interactions limit their use. Therefore, there is an unmet need for direct-acting antiviral agents against the most recent omicron sublineages.

The evaluation of mAbs mostly relies on in vitro neutralization, which limits our understanding of their antiviral properties. Neutralization assays are heterogenous, based on different target cells, and use either viral pseudotypes or authentic isolates^12^. Furthermore, in vitro neutralizing titers do not consider variation in antibody doses and pharmacokinetics, as well as the interaction of mAbs with the endogenous immune response. Antibodies may activate immune cells through interaction between their fragment crystallizable (Fc) and Fc Receptors. These non-neutralizing Fc-effector functions are resilient against viral variants and contribute to therapeutic efficacy in preclinical models^13,14^. Hence, an evaluation of both neutralizing and non-neutralizing activities of mAbs in patient samples is needed to accurately determine their efficacy against SARS-CoV-2 variants.

Here, we longitudinally evaluated neutralization and antibody-dependent cellular cytotoxicity (ADCC) in serum samples of immunocompromised patients receiving sotrovimab, combinations of casirivimab/imdevimab and cilgavimab/tixagevimab, or, as a control, nirmatrelvir. We used authentic viral isolates to determine neutralization titers and a reporter assay to measure ADCC. Our data show that sotrovimab is active against XBB.1.5 and BQ.1.1 upon administration, supporting its use against these variants and suggesting a clinical benefit of increasing its dose.

## Results

Among the 756 patients with mild-to-moderate COVID-19 recruited in the ANRS 0003S CoCoPrev cohort study, we included all immunocompromised individuals with less than 280 BAU/mL of anti-S antibodies at inclusion having a complete longitudinal follow-up (i.e., serum sample available at day 0, day 7, month 1 and month 3; n=80). Among them, 67 were treated with mAbs (sotrovimab, n=29; imdevimab/casirivimab/, n=34; cilgavimab/tixagevimab, n=4), and 13 received nirmaltrelvir/ritonavir. A complete description of patients’ characteristics is available in the **table 1**. Because of the evolution of guidelines and emergence of variants during the study period, treatments and infecting strains changed. Patients treated with imdevimab/casirivimab were mainly infected by Delta, while those who received sotrovimab, cilgavimab/tixagevimab and nirmaltrelvir were mostly infected by Omicron. All patients received at least 1 dose of vaccine, but the proportion of patients who received ≥ 3 doses of vaccine was lower in the imdevimab/casirivimab group as the booster dose was not yet recommended at the time of their inclusion. Age, body mass index and COVID-19 severity were similar across all groups. None of the patients died during the follow-up. All mAbs were used at their indicated doses at the time of inclusion (500mg for sotrovimab, 600mg/600mg or 300mg/300mg for imdevimab/casirivimab and 600mg/600mg for cilgavimab/tixagevimab). Nirmatrelvir was administered for 5 days.

**Table 1.** Characteristics of patients.

|  | **Total**<br>**N=80** | **Casirivimab/ Imdevimab**<br>**(1200 and 600 mg) N=34** | **Sotrovimab**<br>**N=29** | **Nirmatrelvir/r**<br>**N=13** | **Tixagévimab/ Cilgavimab**<br>**N= 4** | **p-value** |
| --- | --- | --- | --- | --- | --- | --- |
| **SARS-CoV-2 variant (n, %)** |  |  |  |  |  | **<0.001** |
| Delta | 33 (42%) | 33 (100%) | 0 | 0 | 0 |  |
| Omicron | 45 (58%) | 0 | 29 (100%) | 12 (92%) | 4 (100%) |  |
| Omicron BA.1 | 27 (33%) | 0 | 26 (90%) | 1 (8%) | 0 | **<0.001** |
| Omicron BA.2 | 14 (17%) | 0 | 3 (10%) | 11 (84%) | 0 |  |
| Omicron BA.5 | 4 (5%) | 0 | 0 | 0 | 4 (100%) |  |
| Missing | 2 | 1 | 0 | 1 | 0 |  |
| **Age (years, median, Q1-Q3)** | 59 (45-70) | 54 (45-64) | 64.5 (42.5-70.5) | 65 (55-73) | 47.5 (32 – 62.5) | 0.16 |
| ≥ 80 years old | 6 (8%) | 3 (9%) | 2 (7%) | 1 (8%) | 0 | 1.0 |
| Missing | 1 | 0 | 1 | 0 | 0 |  |
| **BMI (median, Q1-Q3)** | 25 (23-28) | 26 (21.5-30.5) | 24 (23-28) | 25 (22-27.5) | 25 (22 - 31) | 0.96 |
| Missing | 9 | 6 | 2 | 1 | 0 |  |
| **Male sex (n, %)** | 37 (46%) | 16 (47%) | 14 (48%) | 7 (54%) | 0 | **0.003** |
| **Immunocompromised patients (n, %)** | 80 (100%) | 34 (100%) | 29 (100%) | 13 (100%) | 4 (100%) | - |
| Ongoing chemotherapy | 26 (33%) | 8 (24%) | 9 (31%) | 9 (69%) | 0 | **0.01** |
| Solid organ transplantation | 23 (29%) | 9 (26%) | 12 (41%) | 1 (8%) | 1 (25 %) | 0.14 |
| Hematopoietic stem cell transplant | 3 (4%) | 0 | 2 (7%) | 1 (8%) | 0 | 0.31 |
| Corticosteroids | 9 (11%) | 7 (21%) | 2 (7%) | 0 | 0 | 0.19 |
| Systemic lupus or vasculitis with immunosuppressive medications | 3 (4%) | 1 (3%) | 2 (7%) | 0 | 0 | 0.80 |
| Immunosuppressive therapy including rituximab | 38 (48%) | 18 (53%) | 15 (52%) | 2 (15%) | 3 (75 %) | 0.05 |
| Cancer | 7 (9%) | 0 | 5 (17%) | 2 (15%) | 0 | **0.04** |
| **Other non-immunosuppressive comorbidities (n, %)** | 27 (34%) | 13 (38%) | 10 (34%) | 2 (15%) | 2 (50 %) | 0.41 |
| **Covid-19 severity (n, %)** |  |  |  |  |  | 0.92 |
| Mild | 70 (90%) | 31 (91%) | 24 (86%) | 11 (91%) | 4 (100 %) |  |
| Moderate | 8 (10%) | 3 (9%) | 4 (14%) | 1 (8%) | 0 |  |
| Missing | 2 | 0 | 1 | 1 | 0 |  |
| **Median time between symptoms onset and initiation of treatment (days, median, Q1-Q3)** | 3 (2-4) | 3 (2-4) | 3 (2-4) | 2.5 (1-4) | 3 (2 – 6) | 1.0 |
| Missing | 7 | 1 | 2 | 3 | 1 |  |
| **Vaccination status (n, %)** |  |  |  |  |  | **0.02** |
| Having received ≥ 3 doses of vaccine | 61 (76%) | 20 (60%) | 25 (86%) | 12 (92%) | 4 (100 %) |  |
| Having received ≤ 2 doses of vaccine | 19 (24%) | 14 (40%) | 4 (14%) | 1 (8%) | 0 |  |
| Median time between last vaccine shot and initiation of treatment (days, median, Q1-Q3) | 122.5 (64.5-175) | 132 (67-155) | 126.5 (54-234) | 102.5 (73-126) | 209(107-271) | 0.32 |
| Missing | 8 | 3 | 3 | 1 | 1 |  |
| **Covid-19-related hospitalization at day 28 (%; 95% Clopper-Pearson CI)** | 2 (3, 0-9) | 0 | 2 (7, 1-23) | 0 | 0 | 0.37 |

First, we measured the levels of anti-S IgGs in sera prior to (day 0) or after treatment initiation, at day 7, month 1 and month 3 **(Figure 1)**. We used our standardized S-Flow assay to determine BAU/mL^15^. Overall, patients treated with nirmaltrelvir do not display a notable increase in anti-spike antibody levels during the follow up (medians of 34 [3-50.5], 36 [4.5-107], 40 [5.5 – 1709] and 45 [15 – 1563], at day 0, day 7, month 1 and month 3, respectively). Four out of the 13 patients end the follow-up with BAU/mL above 280 **(Figure 1)**, indicative of an endogenous immune response in the less immunosuppressed individuals. Administration of mAbs leads to a sharp increase of BAU/mL between day 0 and day 7 in all patients (4 [1-31.5] vs. 884 [546-1,309], p<0.0001; 13 [2-34] vs. 1,746 [977-3,522], p<0.0001; 14.5 [2.5-131.5] vs. 3,003 [2,582-4,982], ns; for sotrovimab, imdevimab/casirivimab and cilgavimab/tixagevimab, respectively). Peak values are reached at day 7 for sotrovimab-treated individuals and M1 for those treated with imdevimab/casirivimab and cilgavimab/tixagevimab **(Figure 1)**. At the peak, antibody levels are lower in the sotrovimab groups as compared to imdevimab/casirivimab and cilgavimab/tixagevimab groups (884 [546-1,309] vs. 3037 [664-5,611], p<0.0001; 884 [546-1,309] vs. 3,880 [3,788-26,273] p=0.012; respectively). Overall, these results show that the anamnestic response to S is, as expected, impaired in our study population, but that mAb administration rescues anti-S antibody levels in the serum.

**Figure 1:**
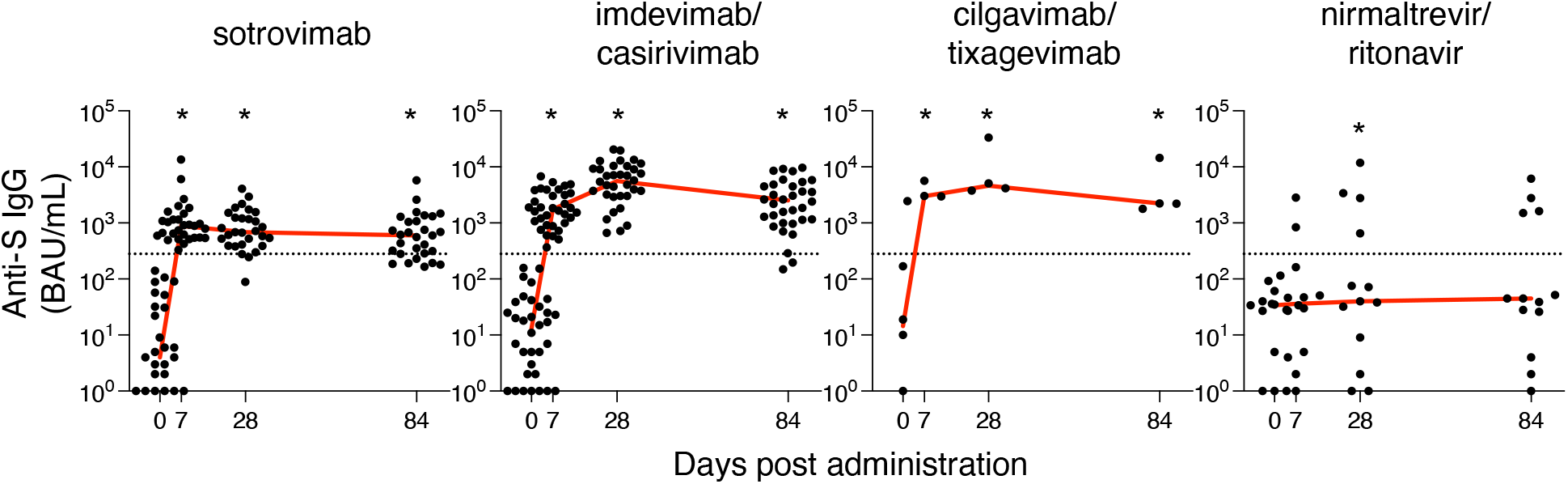
Antibody levels in sera of COVID-19 patients receiving antiviral therapies. (A) Anti-S IgGs were measured using the flow cytometry-based S-Flow assay in sera of individuals before and after treatment initiation. sotrovimab, n=29; imdevimab/casirivimab, n=34; tixagevimab/cilgavimab, n=4; nirmatrelvir/ritonavir, n=13. Indicated are the binding antibody units (BAUs) per mL (BAU/mL) of anti-S IgGs. Stars indicate statistical significance according to a two-sided Friedman test with a Dunn’s multiple comparison correction. Each dot is an individual. Red lines indicate medians. Dashed lines indicate 280 BAU/mL.

Next, we measured the neutralizing activity of these sera with authentic viral isolates of D614G, BQ.1.1 and XBB.1.5 in our S-Fuse assay^16,17^. Administration of mAbs elicited serum neutralization of D614G, which remains detectable during the entire follow-up **(Figure 2 and supplementary figure 1)**. Median titers at day 7 were lower in individuals treated with sotrovimab as compared to those treated with imdevimab/casirivimab or cilgavimab/tixagevimab (1,115 [799-1,674] vs. 100,000[100,000-100,000], p<0.0001; 1,115 [799-1,674] vs. 90,644 [79,516-100,000], ns; respectively). This difference remains through the follow-up. Nirmaltrelvir therapy leads to a non-significant increase in neutralizing titers against D614G, which are, however, highly heterogeneous. Interestingly, this heterogenous antiviral activity in sera of nirmaltrelvir recipients is also observed against BQ.1.1 and XBB.1.5. This may be due to circulating levels of nirmaltrevir that inhibit viral infection in our assay or to the induction of endogenous neutralizing antibodies in some individuals. BQ.1.1 and XBB.1.5 omicron sub-variants dramatically reduce the neutralization provided by mAbs administration. Neither imdevimab/casirivimab nor cilgavimab/tixagevimab administration elicit serum neutralization of BQ.1.1 or XBB.1.5. Some individuals harbor low neutralizing titers above our threshold of 10 at day 7, month 1 and month 3, but medians lack significance when compared to day 0. For these 2 combinations of mAbs, fold decreases for the neutralization of BQ.1.1 or XBB.1.5 as compared to D614G at day 7 are >20,000. Neutralization is also reduced against XBB.1.5 and BQ.1.1 in individuals treated with sotrovimab, albeit to a lower extent. Low levels of neutralization of BQ1.1 and XBB.1.5 are detectable in most sotrovimab-treated individuals, and reached significance as compared to day 0 at month 1 for BQ.1.1 (p=0.008) and at all time points for XBB.1.5 (p=0.0006, p<0.0001, p=0.0003). At day 7, BQ.1.1 and XBB.1.5 neutralizing titers are reduced by 71- and 58-fold as compared to D614G (15.6 [5-24.94] and 18.9 [13.6-56.12] vs. 1,115 [799-1,674], respectively). These results are in line with in vitro evaluation of mAbs against D614G, BQ.1.1 and XBB.1.5, where only sotrovimab remains active against these variants **(Supplementary Figure 2)**. Altogether, these results show that mAb infusion elicits a robust neutralization of SARS-CoV-2 in the serum of treated individuals, which is evaded by BQ.1.1 and XBB.1.5 but remains detectable in individuals treated with sotrovimab.

**Figure 2:**
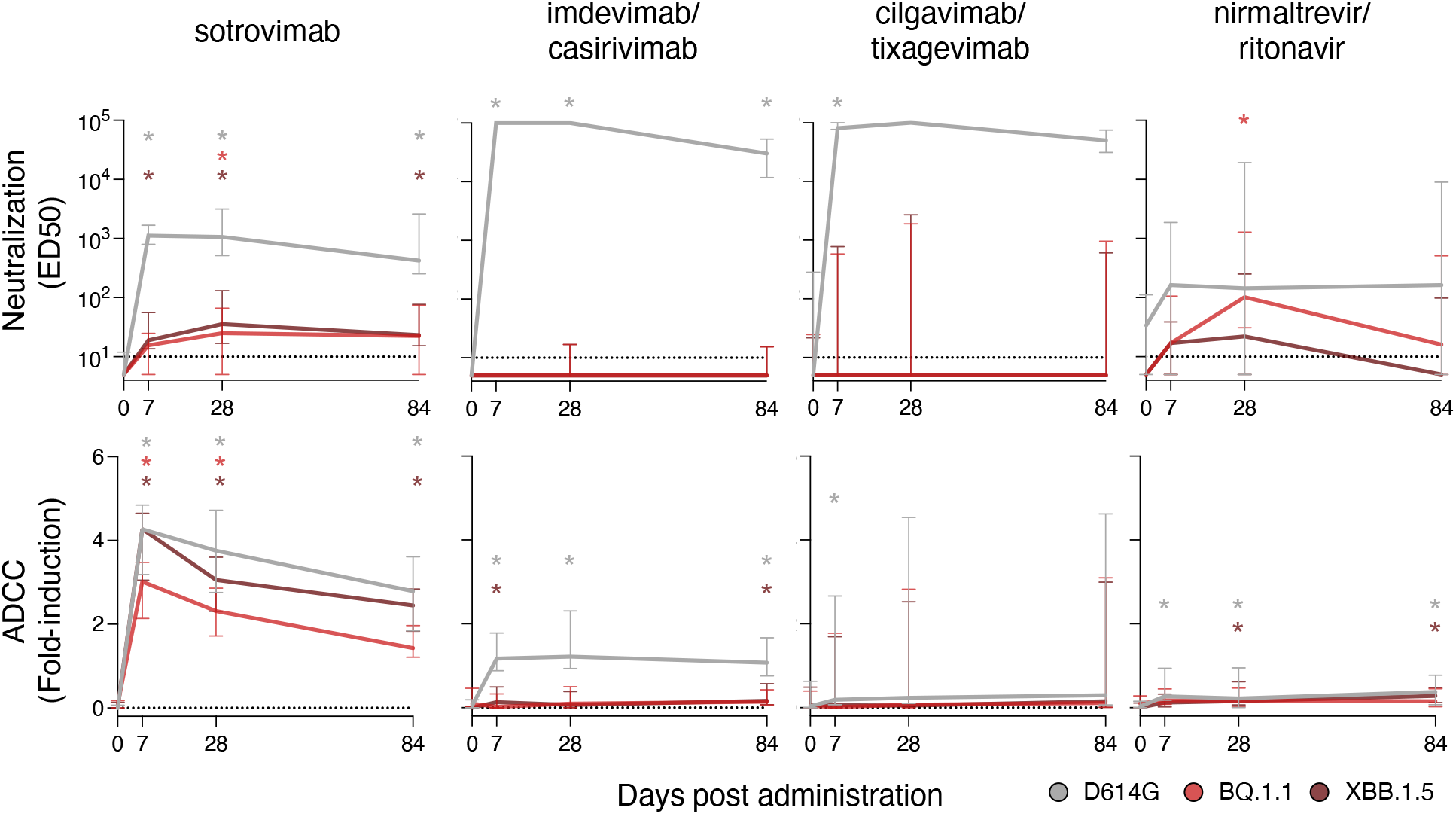
Neutralization and antibody-dependent cellular cytotoxicity of D614G, BQ.1.1 and XBB.1.5 in sera of COVID-19 patients receiving antiviral therapies. Serum neutralization (A) and ADCC activity (B) of D614G and Omicron BQ.1.1 and XBB.1.5 in sera of individuals before and after treatment initiation. sotrovimab, n=29; imdevimab/casirivimab, n=34; tixagevimab/cilgavimab, n=4; nirmatrelvir/ritonavir, n=13. Indicated are either effective dilution 50% (ED50; titers) as calculated with the S-Fuse assay (A) or fold-induction of the CD16 pathway in the ADCC reporter assay (B). Stars indicate statistical significance according to a two-sided Friedman test with a Dunn’s multiple comparison correction. Colored lines indicate medians and error bars inter-quartile range. The dashed lines indicate the limits of detection. Individual data points are depicted in supplementary figure 1.

Finally, we tested the ADCC activity of these sera **(Figure 2 and supplementary figure 1)**. We used our reporter assay based on spike-expressing 293T cells and jurkat cells expressing CD16 and a NFAT-luciferase cassette. We have previously demonstrated that this assay correlates with the ADCC of SARS-CoV-2-infected cells by primary NK cells^18^. Consistent with the evaluation of ADCC in vitro **(Supplementary Figure 1)**, only sotrovimab and imdevimab/casirivimab elicit significant and substantial levels of ADCC against the D614G spike. When measuring ADCC against BQ1.1 and XBB.1.5, only sotrovimab-treated individuals remain active. Of note, fold changes were limited as compared to D614G and only significantly decreased for BQ1.1 (at day 7: 3 [2.1-3.5] vs. 4.3 [3.2-4.8], 1.4-fold, p=0.0075 and 4.3 [3.1-4.6] vs. 4.3 [3.2-4.8], 1-fold, ns, respectively), again in line with the in vitro evaluation **(Supplementary Figure 1)**. Cilgavimab and tixagevimab are mutated in their Fc domain to decrease interactions with FcRs, which likely explained the complete absence of ADCC of this combination even against D614G. Nirmaltrelvir treated individuals exhibit a low and sporadically significant increase in ADCC activity, likely reflecting an endogenous antibody response. Overall, these data show that sotrovimab elicits the strongest level of ADCC activity in serum, as compared to other mAbs. This ADCC activity is only slightly evaded by BQ.1.1 and preserved against XBB.1.5.

## Discussion

Here, we show that sotrovimab neutralizes and elicits ADCC against the omicron sub-variants BQ.1.1 and XBB.1.5. In cell culture systems, we demonstrated that neutralization and ADCC of sotrovimab are reduced but not abrogated against these variants. This is in contrast with imdevimab/casirivimab and cilgavimab/tixagevimab, which lose all antiviral activities even at high concentration. In patient samples, we show that administration of 500 mg of sotrovimab elicits serum neutralization and ADCC of BQ.1.1 and XBB.1.5 for up to 3 months. Administration of imdevimab/casirivimab or cilgavimab/tixagevimab triggers neutralization exclusively against D614G with only limited or undetectable levels of ADCC. As a control, we included individuals treated with nirmaltrelvir, a therapy recommended against BQ.1.1 and XBB.1.5 variants. Its administration led to highly variable, but overall low, levels of viral inhibition in the serum and no ADCC, which can be attributed to either nirmaltrelvir or to low levels of endogenous antibody production.

Our results show that sotrovimab infusion elicits an antiviral immunity against BQ.1.1 and XBB.1.5 in the sera of treated patients. This finding explains why several studies suggested a preserved efficacy of sotrovimab in real-life settings despite its limited neutralization^19–24^. This is consistent with the recent demonstration that very low levels of neutralization are protective upon infusion of a polyfunctional mAbs in human^25^. It is also in line with the capacity of sotrovimab and its precursor S309 to limit viral replication in preclinical models of BQ.1.1 infection^26,27^. S309 relies on both neutralization and Fc-effector functions to limit SARS-CoV-2 replication in preclinical models^28^. Further work is needed to determine the contribution of neutralizing vs non-neutralizing activities to the clinical efficacy of sotrovimab.

The level of BAU/mL measured in the serum is lower in individuals receiving sotrovimab than in those treated with imdevimab/casirivimab and cilgavimab/tixagevimab. Both neutralization and ADCC against D614G, BQ.1.1 and XBB.1.5 are correlated to BAU/mL (**Supplementary figure 3 and 4**). In vitro, spiking of sotrovimab in sera of patients with blood cancer increases neutralization in a dose-dependent manner^29^. Thus, increasing sotrovimab dose will likely lead to an increase of its in vivo antiviral activities, as previously reported for cilgavimab/tixagevimab^30^. Given the good safety profile of sotrovimab and of anti-S antibodies in general (up to 4000/4000 mg for imdevimab/casirivimab for instance), increasing sotrovimab dose may increase its clinical value, a hypothesis that is being tested at 1000 mg in the RECOVERY clinical trial (NCT04381936). Combination therapy with sotrovimab and antivirals to improve virological and clinical responses in immunocompromised COVID-19 patients also deserves investigation^31^.

In conclusion, our data supports the use of sotrovimab to treat the most recent omicron lineages and suggest that increasing the dose may provide a clinical benefit.

### Limitations of the study

Our study has limitations. First, we do not have a control group of untreated patients because of ethical reasons. We thus included patients treated with nirmaltrelvir as control. Their investigation confirms that our inclusion criteria selected for patients with impaired endogenous immune response to SARS-CoV-2. Some individuals seroconverted, but this is explained by the heterogenous nature of immunocompromised individuals. However, 8 out of the 13 individuals from this control group, which were included with the same criteria, failed to seroconvert. This confirms that most of the effects observed in groups of mAb-treated individuals are linked to the injected antibodies, rather than an endogenous immune response. This is further confirmed by similarity between the findings obtained with the antibody alone and those in the sera of patients. Since endogenous immune responses are negligible in our cohort, the difference in infecting variant is also unlikely to bias our conclusion, albeit we can’t rule out that it influences the results at the individuals’ level. We did not directly quantify the antibodies in the sera and restricted our analysis to BAU/mL. Still, for the same reason, measured BAU/mL mostly quantified the injected antibody. Our neutralization assay may be criticized because of the use of ACE2-overexpressing cells. Yet, ACE2 overexpression underestimates sotrovimab efficacy^12,32^. Therefore, using systems with endogenous ACE2 expression, such as Vero cells would lead to higher neutralization titers, further confirming our conclusion. Our ADCC assay is a surrogate assay measuring CD16 activation rather than cell death by NK cells. However, we have previously demonstrated that it correlates with an authentic ADCC assay, using infected cells and primary NK cells^18^. Furthermore, standardization of NK-based ADCC assay is challenging on a large cohort as presented here. Finally, we are not exhaustive in the evaluation of antivirals, as no patient treated with remdesivir, molnupinavir or bebtelovimab were included in our study, the latter two because of the lack of European Medical Agency (EMA) approvals.

## Methods

### Patient Samples

Patient samples were obtained from the ongoing multicentre prospective observational cohort study ANRS 0003S CoCoPrev (Prevention of COVID-19 complications in high-risk SARS-CoV-2-infected subjects eligible for treatments under an emergency use authorization or early access, NCT04885452, ^33^) This cohort recruited consecutively patients at high-risk for progression to severe COVID-19, having PCR-proven mild-to-moderate COVID-19 in the first 5 days of symptoms and who were treated under an emergency use authorization or early access in one of the 32 participating centers. Here, we included vaccinated immunocompromised patients having IgG anti-spike sera titers less than 280 BAU/mL at inclusion and having a serum sample collected at day 0, day 7, month 1 and month 3 after treatment initiation. Blood samples were centralized and stored at “Centre de Ressources Biologiques ANRS-MIE, Bordeaux” and then extracted to the virological centralized unit (Pitié Salpêtrière, APHP, Paris) for further analyses. The protocol was approved by the “CPP Sud-Est IV” Ethics Committee (Paris, France) and the French Regulatory Authority. Written informed consent was obtained from each patient before enrolment.

### Viral isolates

The reference D614G and BQ.1.1 isolates were previously described^34^. The XBB.1.5 strain was isolated on IGROV-1 cells from a nasopharyngeal swab of an anonymous individual attending the emergency room of Hôpital Européen Georges Pompidou (HEGP; Assistance Publique, Hôpitaux de Paris). Sequencing of the nasopharyngeal swab, and the isolated virus confirmed XBB.1.5 lineage. For sequencing, we used an untargeted metagenomic sequencing approach with ribosomal RNA depletion as previously described^34^. The sequences were deposited on GISAID (D614G: EPI_ISL_414631; BQ.1.1: ID: EPI_ISL_15731523; XBB.1.5: EPI_ISL_16353849). Viral stocks were prepared from isolated strains using either Vero E6 cells or IGROV-1 cells and titrated using Vero E6 and S-Fuse cells.

### Neutralization

Serum neutralizing titers were measured using the S-Fuse assay as previously described^35^. Briefly, U2OS-ACE2 GFP1-10 or GFP11 cells, also termed S-Fuse cells, become GFP + when infected by SARS-CoV-2. Cells expressing GFP1-10 or GFP11 were mixed (ratio 1:1) and plated at 2×10^4^ cells per well in a mClear 96-well plate (Greiner Bio-One). Indicated SARS-CoV-2 strains were incubated with serially diluted mAb or sera and added to S-Fuse cells after a 15min incubation at room temperature. All sera were heat-inactivated for 30 minutes at 56°C. Cells were then incubated 18h at 37°C 5%CO_2_, fixed with 4% paraformaldehyde (PFA), washed and stained with Hoechst (dilution 1:10,000, Invitrogen). Images were acquired with an Opera Phenix high-content confocal microscope (PerkinElmer). The GFP area and the number of nuclei were quantified using Harmony software version 4.9 (PerkinElmer). The percentage of neutralization was calculated using the number of syncytia as value with the following formula: 100x(1-(“value with serum”-”value in non-infected”)/(“value in no serum”-”value in non-infected”)). Effective dose 50% (ED50, aka “titers”), in mg.ml^-1^ for mAbs and in dilution values for sera, were calculated with a reconstructed curve using the percentage of the neutralization at the different concentrations. We previously reported correlations between neutralization titers obtained with the S-Fuse assay and both pseudovirus neutralization and microneutralization assays^36,37^.

### Antibody-dependent cellular cytotoxicity

ADCC was quantified using the ADCC Reporter Bioassay (Promega). Briefly, 293T cells were transfected with plasmids (pVAX1, Invitrogen) encoding the indicated spikes using lipofectamine 2000 (ThermoFisher) and incubated overnight to allow for transgene expression. All spike sequences were codon-optimized, and the cytoplasmic tail was deleted (last 19 Amino acids). The day of the assay, transfected cells (3×10^4^ per well) were co-cultured with Jurkat-CD16-NFAT-rLuc cells (3×10^4^ per well) in presence or absence of mAbs at the indicated concentration. The expression of spikes was controlled by flow cytometry using the previously described pan-coronavirus antibody (mAb10)^38^. Luciferase was measured after 18 h of incubation using an EnSpire plate reader (PerkinElmer). ADCC was measured as the fold induction of Luciferase activity compared to the ‘‘no serum’’ condition. Sera were tested at a 1:30 dilution. For each serum, the control condition (cells transfected with an empty plasmid) was subtracted to account for inter-individual variations of the background. We previously reported correlations between the ADCC Reporter Bioassay titers and an ADCC assay based on primary NK cells and cells infected with an authentic virus^18^.

### Statistical analysis

No statistical methods were used to predetermine sample size. The experiments were not randomized, and the investigators were not blinded. Calculations were performed using Excel 365 (Microsoft). Figures and statistical analysis were performed using GraphPad Prism 9 (GraphPad Software). Unless indicated, median and inter-quartile range are use to describe the data. Statistical significance between different groups was calculated using Kruskall–Wallis test with Dunn’s multiple comparisons, Friedman test with Dunn’s multiple comparison correction and Spearman non-parametric correlation test. All tests were two-sided.

## Data Availability

All data produced in the present study are available upon reasonable request to the authors.

## Acknowledgement

We thank members of the Virus and Immunity Unit for discussions and help in the preparation of this manuscript. We thank Valentin Leducq and Emna Ghidaoui for their technical assistance. We thank Laurent hocqueloux and Thierry Prazuck for providing the clinical antibodies for in vitro studies and Faustine Amara for her help with the plasmids. We thank Pr Yazdan Yazdanpanah and all the ANRS-MIE team for their invaluable support and help. This study would have not been possible without the teams involved in the CoCoPrev Study and designated as the CoCoPrev Study Group: Magali Garcia, Valentin Giraud, Agathe Metais, France Cazenave-Roblot, Jean-Philippe Martellosio (CHU de Poitiers); Anne-Marie Ronchetti, Thomas Gabas, Naima Hadjadj, Célia Salanoubat, Amélie Chabrol, Pierre Housset, Agathe Pardon, Anne-Laure Faucon, Valérie Caudwell, Latifa Hanafi (CHU Sud Francilien, Corbeil-Essonne) ; Laurent Alric, Grégory Pugnet, Morgane Mourguet, Eva Bories, Delphine Bonnet, Sandrine Charpentier, Pierre Delobel, Alexa Debard, Colleen Beck, Xavier Boumaza, Stella Rousset (CHU de Toulouse) ; Fanny Lanternier, Claire Delage, Elisabete Gomes Pires, Morgane Cheminant, Nathalie Chavarot (Hôpital Necker, Paris) ; Anthony Chauvin, Xavier Eyer ; Véronique Delcey (Hôpital Lariboisière, Paris) ; Simon Bessis, Mélanie Cresta, Romain Gueneau (Hôpital du Kremlin Bicêtre) ; Pelagie Thibaut, Marine Nadal, Martin Siguier, Marwa Bachir, Christia Palacios (Hôpital Tenon, Paris) ; Valérie Pourcher, Cléa Melenotte, Antoine Faycal, Vincent Berot, Cécile Brin, Siham Djebara, Karen Zafilaza, Stéphane Marot, Sophie Sayon, Valentin Leducq, Isabelle Malet, Elisa Teyssou, Adélie Gothland (Hôpital de la Pitié Salpétrière, Paris) ; Karine Lacombe, Yasmine Abi Aad, Thibault Chiarabini, Raynald Feliho, Nadia Valin, Fabien Brigant, Julien Boize, Pierre-Clément Thiébaud, Marie Moreau, Charlotte Billard (Hôpital St Antoine, Paris), Nathalie De Castro, Geoffroy Liégeon, Blandine Denis, Jean-Michel Molina, Lucia Etheve (Hôpital Saint Louis, Paris) ; André Cabié, Sylvie Abel, Ornella Cabras, Karine Guitteaud, Sandrine Pierre-François (CHU de Martinique) ; Vincent Dubee, Diama Ndiaye, Jonathan Pehlivan, Michael Phelippeau, Rafael Mahieu (CHU d’Angers) ; Alexandre Duvignaud, Thierry Piston, Arnaud Desclaux, Didier Neau, Charles Cazanave (CHU de Bordeaux) ; Jean-François Faucher, Benjamin Festou, Magali Dupuy-Grasset, Véronique Loustaud-Ratti, Delphine Chainier (CHU de Limoges) ; Nathan Peiffer-Smadja, Christophe Choquet, Olivia Da Conceicao, Michael Thy, Lio Collas, Cindy Godard, Donia Bouzid, Vittiaroat Ing, Laurent Pereira, Thomas Pavlowsky, Camille Ravaut (Hôpital Bichat, Paris) ; Antoine Asquier-Khati, David Boutoille, Marie Chauveau, Colin Deschanvres, François Raffi (CHU de Nantes) ; Audrey Le Bot, Marine Cailleaux, François Benezit, Anne Maillard, Benoit Hue, Pierre Tattevin (CHU de Rennes) ; François Coustilleres, Claudia Carvalho-Schneider, Simon Jamard, Laetitia Petit, Karl Stefic (CHU de Tours) ; Natacha Mrozek, Clement Theis, Magali Vidal, Leo Sauvat, Delphine Martineau (CHU de Clermond-Ferrand) ; Benjamin Lefèvre, Guillaume Baronnet, Agnès Didier (CHRU de Nancy) ; Florence Ader, Thomas Perpoint, Anne Conrad, Paul Chabert, Pierre Chauvelot (CHU de Lyon) ; Aurélie Martin, Paul Loubet, Julien Mazet, Romaric Larcher, Didier Laureillard (CHU de Nîmes) ; Mathilde Devaux (Hôpital de Poissy); Jérôme Frey, Amos Woerlen, Aline Remillon, Laure Absensur-Vuillaume, Pauline Bouquet (CHU de Metz) ; Albert Trinh-Duc, Patrick Rispal (Hôpital d’Agen) ; Philippe Petua, Julien Carillo (Hôpital de Tarbes) ; Aurore Perrot, Karen Delavigne, Pierre Cougoul, Jérémie Dion, Odile Rauzy (Oncopole, Toulouse), Mathieu Blot, Thibault Sixt, Florian Moretto, Carole Charles, Lionel Piroth (CHU de Dijon) ; Sophie Circosta, Lydia Leger, Arulvani Arulananthan, Carine Lascoux, Pascaline Valérie, Léia Becam (Team Biobanque ANRS-INSERM US19,Villejuif) ; Yazdan Yazdanpanah, Ventzislava Petrov-Sanchez, Alpha Diallo, Soizic Le Mestre, Guillaume Le Meut (ANRS-MIE) ; Isabelle Goderel, Frédéric Chau, Brahim Soltana, Jessica Chane Tang (IPLESP), Jeremie Guedj (Université de Paris, IAME, INSERM, Paris), Yvanie Caille (Renaloo).

## Author contributions

Experimental strategy and design: T.B., O.S., A.G.M. & G.M.B

Laboratory experiments: T.B., L.L.V, F.P, I.S, D.P, F.G., O.S.

Cohort management and clinical research: C.S., K.Z., C.L.N, M.L.M., C.D., D.M., Y.Y., F. C., A.G.M., G.M.B., & C.S.G

Viral strains and key reagents: J.P., M.P., S.M., H.P., E.S.L., & D.V.

Manuscript writing & editing: T.B., O.S., AGM, GMB

## Data Availability

All data produced in the present study are available upon reasonable request to the authors.

---

**Supplementary Figure 1:**
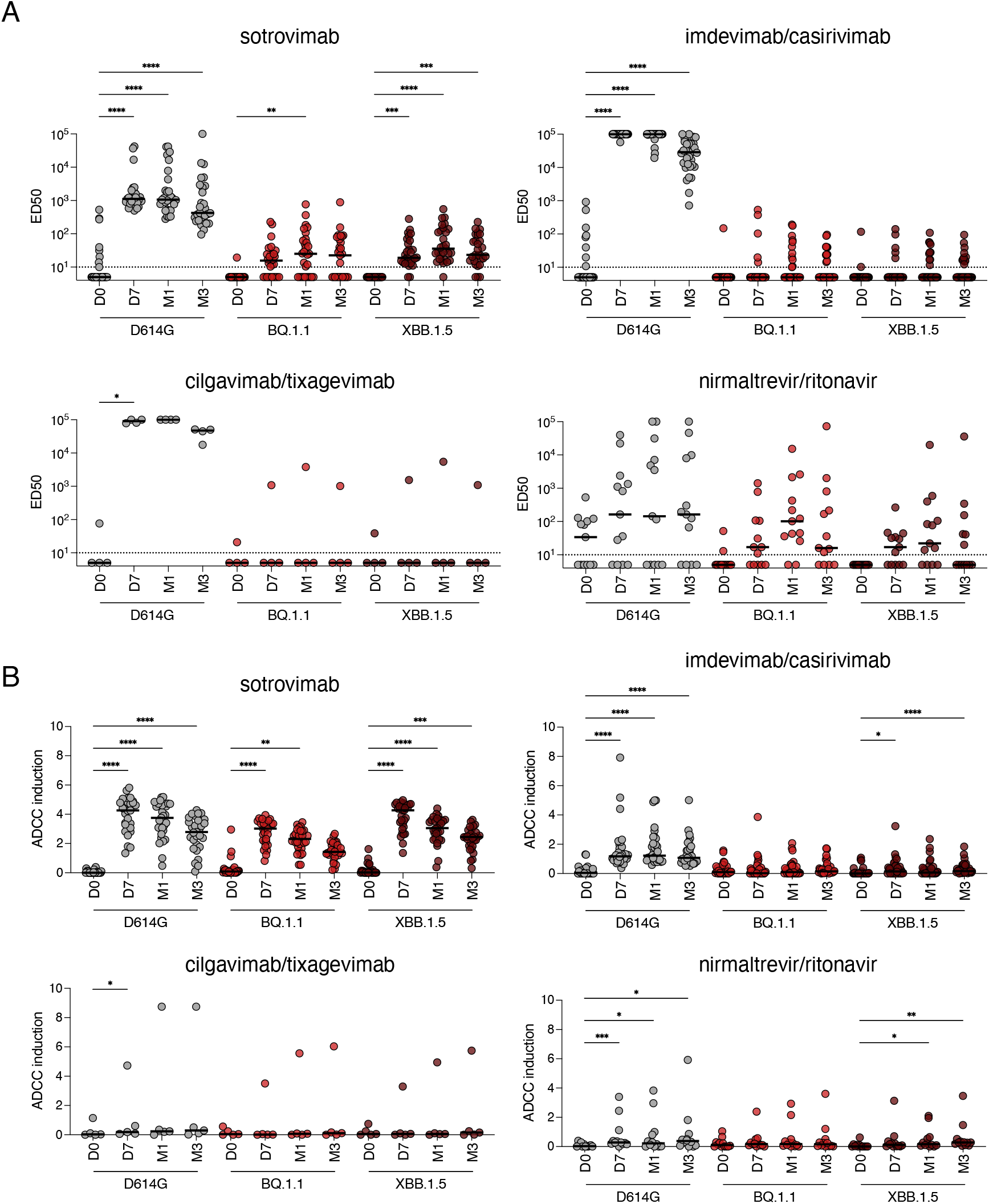
Neutralization and antibody-dependent cellular cytotoxicity of D614G, BQ.1.1 and XBB.1.5 in sera of COVID-19 patients receiving antiviral therapies. Serum neutralization (A) and ADCC activity (B) of D614G and Omicron BQ.1.1 and XBB.1.5 in sera of individuals before and after treatment initiation. sotrovimab, n=29; imdevimab/casirivimab, n=34; tixagevimab/cilgavimab, n=4; nirmatrelvir/ritonavir, n=13. Indicated are either effective dilution 50% (ED50; titers) as calculated with the S-Fuse assay (A) or fold-induction of the CD16 pathway in the ADCC reporter assay (B). Stars indicate statistical significance according to a two-sided Friedman test with a Dunn’s multiple comparison correction. Each dot is an individual. Black lines indicate medians.

**Supplementary Figure 2:**
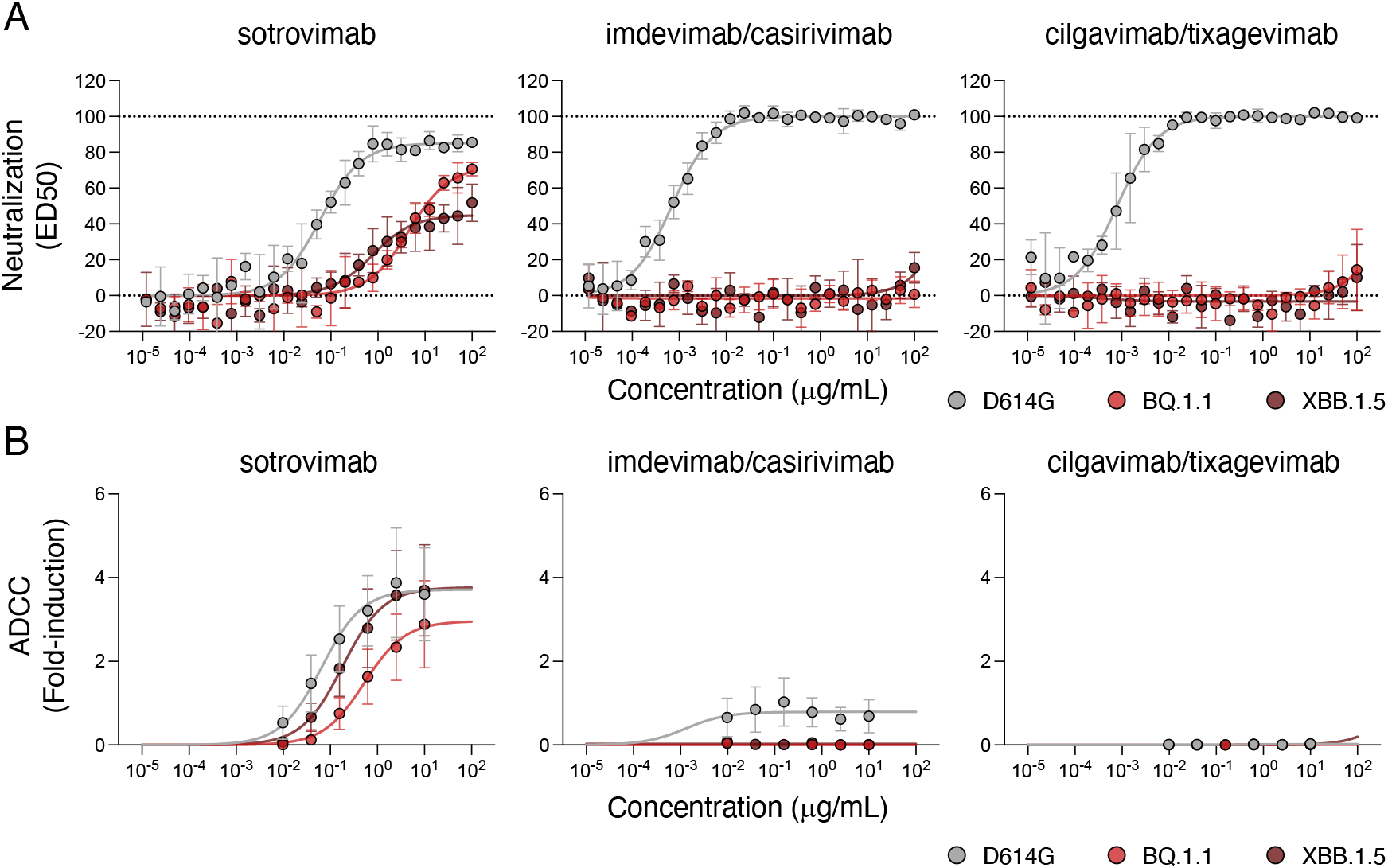
In vitro evaluation of neutralization of antibody-dependent cellular cytotoxicity of COVID-19 therapeutic mAbs against D614G, BQ.1.1 and XBB.1.5. (A) Neutralization curves of mAbs using the S-Fuse system. Dose-response analysis of the neutralization by the indicated antibodies. Data are mean ± SD of 3 independent experiments. The dashed lines indicates no inhibtion (0%) and full inhibition (100%). (B) Dose–response analysis of the ADCC activity by the indicated antibodies. The y-axis indicates the fold-increase in CD16 induction, calculated using the condition without target cells and with target cells transfected with a empty plasmid. Data are mean ± s.d. of 2-5 independent experiments.

**Supplementary Figure 3:**
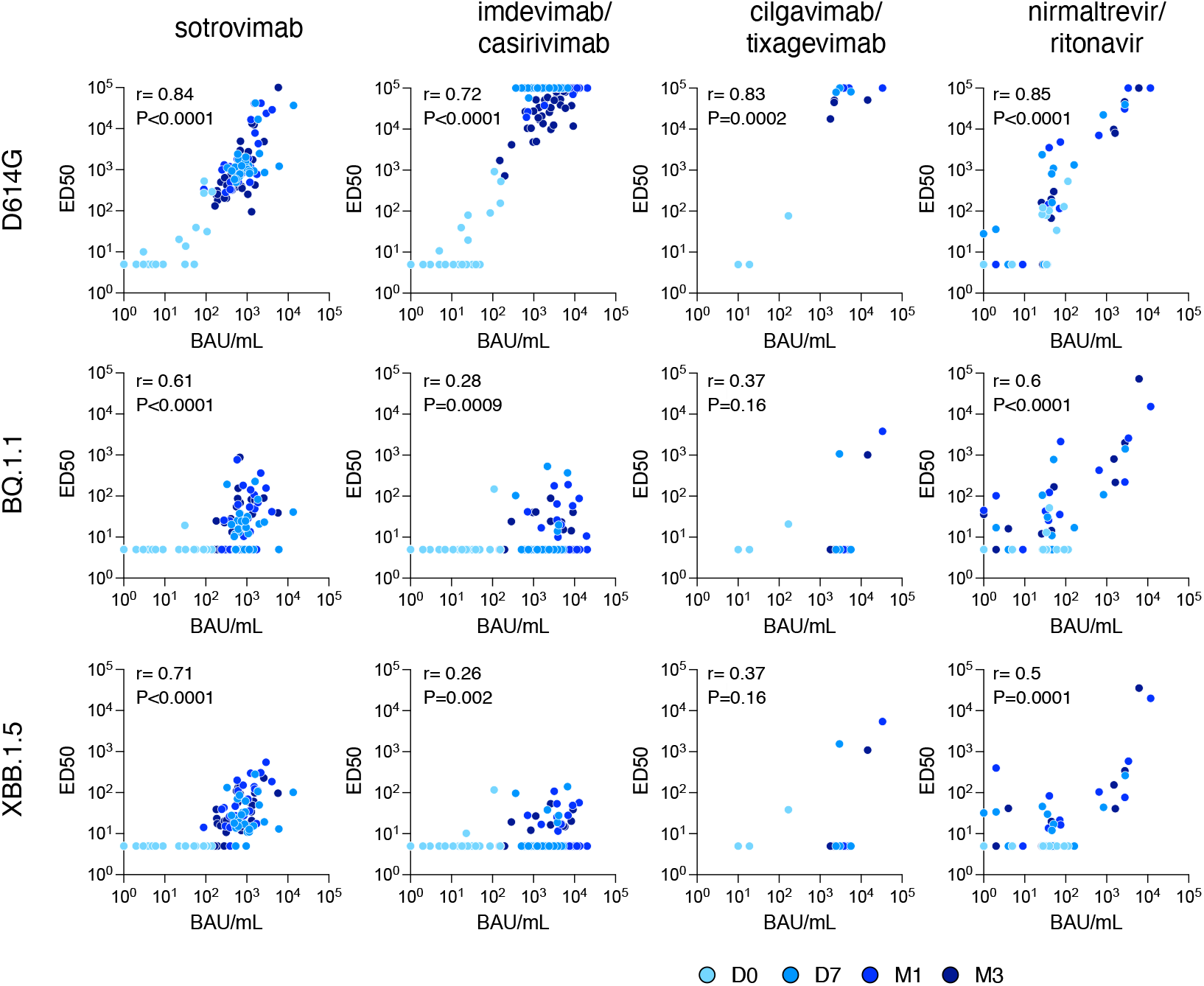
Correlation between serum neutralization of D614G, BQ.1.1 and XBB.1.5 and BAU/mL. Correaltion between BAU/mL and serum neutralization of D614G, BQ.1.1 and XBB.1.5 as measured in the S-Fuse system. Each dot is a patient. Colors indicate days or months post treatment. R and p values are calculated with the Spearman non-parametric correlation test.

**Supplementary Figure 4:**
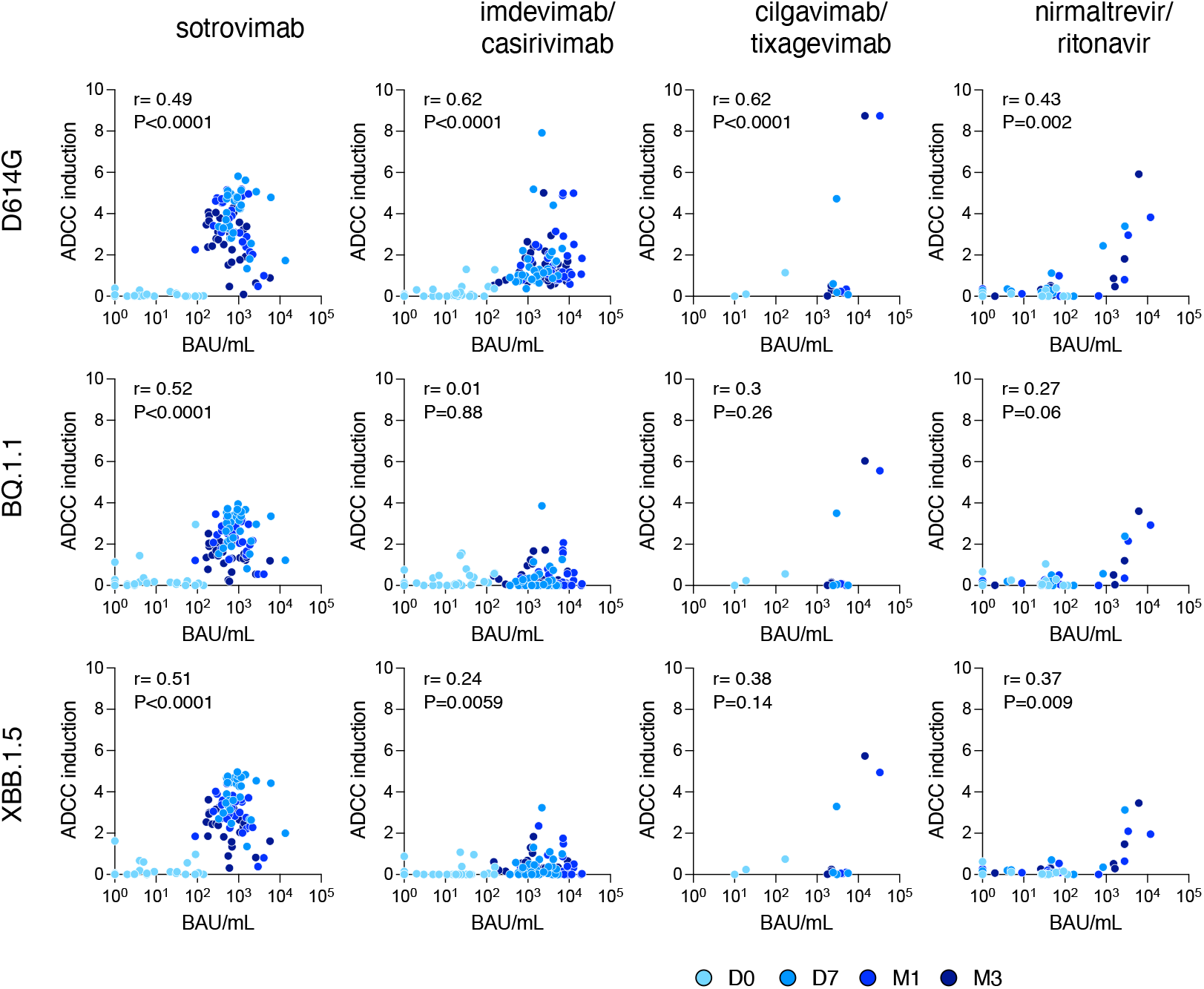
Correlation between serum ADCC of D614G, BQ.1.1 and XBB.1.5 and BAU/mL. Correaltion between BAU/mL and serum ADCC of D614G, BQ.1.1 and XBB.1.5 as measured in the ADCC reporter system. Each dot is a patient. Colors indicate days or months post treatment. R and p values are calculated with the Spearman non-parametric correlation test.

